# A Large Spanish Cohort Study Defines SCA27B Distinct Clinical Phenotype and its Longitudinal Progression

**DOI:** 10.64898/2026.09.11.26362788

**Authors:** Ana Vinagre-Aragón, Gonzalo Olmedo-Saura, Idoia Rouco Axpe, Astrid Daniela Adarmes Gómez, Paula Pérez Torre, Berta Alemany-Perna, Beatriz Castillo Calvo, María Fernanda Pacheco Mendoza, Lorena Garayoa Telletxea, Raquel Baviera-Muñoz, German Morís, Ines Albajar Gómez, Silvia Martí, Sandra Colina, Jorge Alonso Pérez, Valentina Vélez-Santamaría, Jone Bocos Portillo, Leire Manrique, Jose Gazulla, Laura Rojas-Bartolome, Elena Bellosta Diago, Antonio Méndez Guerrero, Almudena Ávila, María Jesús Sobrido, Miguel Ángel Rubio, Daniel López Domínguez, Stéphane Fourcade, Rafael Sivera, Solange Kapetanovic, Izaro Kortazar, Esteban Muñoz, Andrea Manuela Mirea, Cecilia Gil Polo, Virginia García-Solaesa, Claudio Catalli, David Pellerin, Marie-Josée Dicaire, Henry Houlden, Bernard Brais, David Campo-Caballero, Elisabet Mondragón Rezola, Lara Pardina Vilella, María Sofía Cámara Marcos, Roberto Fernández-Torrón, Patricia Garay Albizuri, Lorea Blazquez, Victoria Álvarez, Jon Infante Ceberio, Luis Bataller Alverola, Pablo Mir, Jesús Pérez-Pérez, Carlos Casasnovas, Javier Ruiz Martínez, Aurora Pujol, Adolfo López de Munain, Pilar Camaño González, Pablo Iruzubieta

## Abstract

**Background:** Heterozygous GAA-TTC repeat expansions in the *FGF14* gene cause spinocerebellar ataxia 27B (SCA27B), a late-onset cerebellar ataxia (LOCA) increasingly recognized in populations of European ancestry.

**Objective:** To characterize the clinical and genetic features of SCA27B and compare its phenotype and progression with those of unexplained idiopathic LOCA (ILOCA) in a large multicenter cohort.

**Methods:** We conducted a nationwide multicenter study of patients with unexplained LOCA recruited from 21 Spanish hospitals. FGF14 GAA-TTC repeat expansions were analyzed using a standardized molecular protocol. Clinical, neuroimaging, and longitudinal data were compared between patients with genetically confirmed SCA27B and those with ILOCA. Longitudinal changes in the Scale for the Assessment and Rating of Ataxia (SARA) were analyzed using linear mixed-effects models.

**Results:** Among 430 patients with unexplained LOCA, 111 (25.8%) carried *FGF14* expansions >250 repeats, and 16 additional patients (3.7%) had intermediate expansions of 200–249 repeats. Including 90 externally recruited genetically confirmed cases, the SCA27B cohort comprised 217 patients. Compared with 283 patients with ILOCA, SCA27B patients had a later age at onset (median 63 vs 55 years, p<0.001), more frequent episodic symptoms (45.2% vs 12.7%), and more frequent oculomotor abnormalities (82.5% vs 71.6%). Downbeat nystagmus was particularly frequent in SCA27B (67.4% vs 32.9%). Conversely, extracerebellar features, including pyramidal and sensory signs, were more common in ILOCA. Among patients receiving 4-aminopyridine (4-AP), subjective improvement was reported more frequently in SCA27B than ILOCA (72.5% vs 36.0%). Longitudinal SARA analysis showed similar rates of progression between groups (SCA27B slope 0.37 vs ILOCA 0.35 points/year; interaction p=0.56).

**Conclusions:** SCA27B was a frequent genetic cause of unexplained LOCA in this nationwide Spanish cohort and showed a distinct clinical phenotype characterized by later onset, episodic symptoms, oculomotor abnormalities, and frequent subjective response to 4-AP. Disease progression was similar to that observed in ILOCA. These findings support systematic *FGF14* testing in patients with late-onset cerebellar ataxia, particularly those with characteristic clinical features.

## Introduction

GAA-TTC repeat expansions in intron 1 of the *FGF14* gene cause spinocerebellar ataxia 27B (SCA27B; MIM:620174), an autosomal dominant late-onset cerebellar ataxia (LOCA) (1,2). Accumulating evidence indicates that SCA27B represents one of the most frequent genetic causes of LOCA in populations of European ancestry (1–4).

Pathogenicity is motif and repeat-length dependent. Non-GAA pure repeats are non-pathogenic (5) while GAA-TTC pure expansions are pathogenic depending on repeat size. Alleles with ≥300 repeats are highly penetrant, whereas alleles with 250–299 repeats show reduced penetrance (1,2). More recent studies suggest that the lower pathogenic threshold may be broader than initially proposed, with 200-249 GAA-TTC repeats being also pathogenic in some individuals (6–8).

Despite the growing recognition of SCA27B, several clinically relevant questions remain poorly studied. These include large studies analyzing SCA27B prevalence, comparing the clinical and neuroimaging features that distinguish SCA27B from idiopathic late-onset cerebellar ataxia (ILOCA), the relationship between repeat length and age at onset, a better characterization of the 4-aminopyridine (4-AP) response, and the longitudinal course of the disease (4,9–11)

To investigate the contribution of SCA27B to unexplained LOCA in Spain, we analyzed a large multicenter cohort of 430 patients. Moreover, we leveraged this cohort and analyzed a large cohort of 217 patients with SCA27B, one of the largest genetically confirmed SCA27B series reported to date (4,6,9,10,12), to identify clinical and paraclinical features that facilitate diagnosis, investigate genotype–phenotype correlations, and provide longitudinal data to better characterize disease progression.

## Methods

### Patients and study design

This multicenter study included patients with LOCA, defined as disease onset at ≥25 years, without a previous genetic diagnosis at enrolment (13). Participants were recruited from 21 Spanish tertiary and university-affiliated centers, providing nationwide coverage (Supplementary Figure 1A). Details of previous genetic investigations are provided in Supplementary Table 1.

A total of 430 patients with unexplained LOCA underwent *FGF14* GAA-TTC repeat expansion testing at the Molecular Diagnosis Platform of the Biogipuzkoa Health Research Institute using a previously described protocol (11). Repeat length was defined as the number of pure GAA-TTC repeats. 64 of these patients were previously published (3). For further analyses, 90 additional genetically confirmed Spanish patients with SCA27B from external laboratories were included (Supplementary Figure 1B). 30 of these patients have been previously published (14,15).

Clinical and demographic data were collected using a standardized REDCap database. Patients carrying *FGF14* repeat expansions were compared with non-carriers (ILOCA group). Among expansion carriers, subgroup analyses were performed according to GAA-TTC repeat length (200-249, 250-299, and ≥300 repeats) and allelic status (monoallelic versus biallelic expansions). Both cross-sectional and retrospective longitudinal analyses were performed.

### Statistical analysis

Statistical analyses were performed using R (version 4.5.2). Continuous variables are presented as median (interquartile range [IQR]) and categorical variables as counts (percentages). Group comparisons were performed using Student’s t-test or Mann–Whitney U test for continuous variables and χ² or Fisher’s exact test for categorical variables, as appropriate. Analyses across repeat-length groups were performed using the Kruskal–Wallis test for continuous variables and χ² or Fisher’s exact test for categorical variables, with false discovery rate (FDR) correction for multiple comparisons where appropriate.

Intergenerational repeat-length variability was assessed in available parent–offspring pairs with parental and offspring allele sizes available (n = 12 pairs). Repeat-length change was calculated as the difference between offspring and parental repeat size (offspring allele size − parental allele size), with positive values indicating expansion and negative values indicating contraction. The association between parental and offspring repeat size was initially assessed using a linear regression model including parental allele size as the sole predictor. A second model additionally included parental sex, and a third model included the interaction between parental allele size and parental sex to assess sex-specific effects on repeat transmission. Nested models were compared using analysis of variance, and changes in explained variance were assessed using the R² and adjusted R² values. In the interaction model, sex-specific regression slopes were derived to quantify the relationship between parental and offspring repeat size separately for maternal and paternal transmissions. Given the limited number of informative parent–offspring pairs, these analyses were considered exploratory.

Longitudinal SARA progression was analyzed using linear mixed-effects models fitted by restricted maximum likelihood (REML), with participant-specific random intercepts to account for repeated measurements. Disease duration (years from symptom onset) was included as a continuous fixed effect in all models, and all models were adjusted for age at evaluation. To compare disease progression between SCA27B and ILOCA, models included diagnosis, disease duration, and their interaction as fixed effects. Additional models assessed the effects of sex and 4-AP treatment on disease progression among patients with SCA27B. The analysis of sex was performed in the overall SCA27B cohort and repeated as a sensitivity analysis restricted to patients with two or more study visits. A sensitivity analysis of 4-AP treatment was restricted to patients who changed treatment status during follow-up, allowing within-patient comparisons. Regression coefficients (β) with 95% confidence intervals are reported for mixed-effects models. All tests were two-sided, and p < 0.05 was considered statistically significant.

### Ethics

All participants provided written informed consent before inclusion and genetic testing. The study was approved by the Basque Country Clinical Research Ethics Committee (CEIm-E; PI2023093) and ratified by the local ethics committees of participating centers.

## Results

### Prevalence of SCA27B in a large cohort of Spanish patients with LOCA

A total of 430 patients with unexplained LOCA were screened for FGF14 GAA-TTC repeat expansions. Pathogenic expansions (>250 GAA-TTC repeats) were identified in 111 patients (25.8%), including 90 (20.9%) carrying ≥300 repeats and 21 (4.9%) carrying 250–299 repeats. Sixteen additional patients (3.7%) harbored expansions of 200–249 GAA-TTC repeats (Figure 1A).

**Figure 1.**
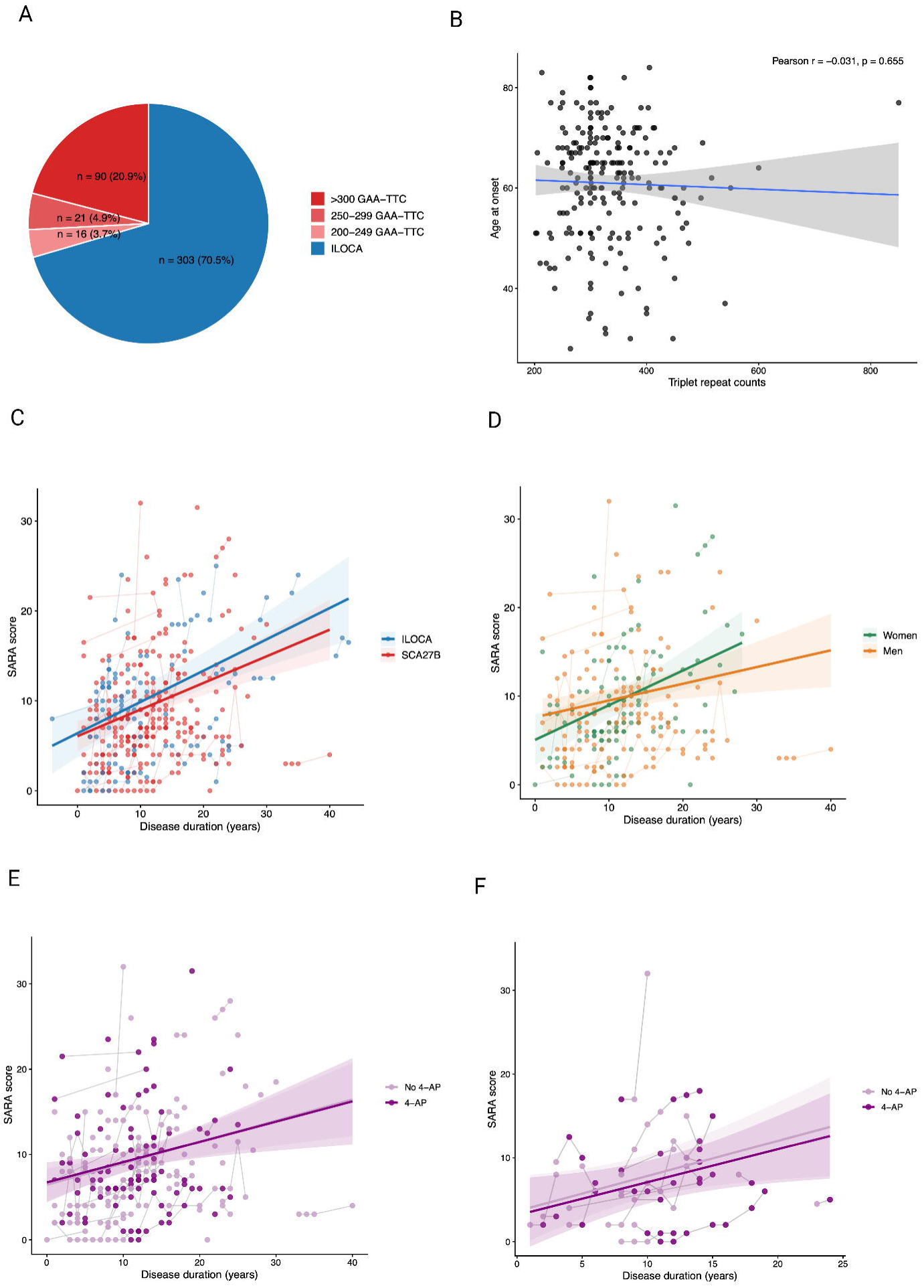
(A) SCA27B prevalence and repeat expansion size distribution. (B) Repeat length was not associated with age at onset (p = 0.65). (C) Longitudinal SARA progression by diagnosis. (D) By sex. (E) By 4-AP treatment. (F) In patients changing 4-AP status.

### Differential features between patients with SCA27B and ILOCA

We then compared the main clinical features of patients with SCA27B with those of patients with ILOCA. For these analyses, we added to the previous cohort 90 genetically confirmed SCA27B cases from external Spanish laboratories tested using the same previously described protocol (11) (n=217 patients with >200 GAA-TTC repeats), yielding one of the largest SCA27B cohorts assembled to date (Supplementary Figure 1B).

Patients with SCA27B had a significantly later age at onset than those with ILOCA (median, 63 vs 55 years; p-FDR < 0.001; Supplementary Figure 2) and were older at evaluation (74 vs 66 years; p-FDR < 0.001; Table 1). Disease duration at evaluation was similar between groups (10 vs 9 years; p-FDR = 0.251). Ataxia severity did not differ significantly between groups, as assessed by SARA (8.5 vs 10; p-FDR = 0.390) or Spinocerebellar Degeneration Functional Score (SDFS) (4 vs 3; p-FDR = 0.751). Similarly, disease duration at SARA and SDFS assessment and corresponding rates did not differ significantly between groups (all p-FDR ≥ 0.751). Age at SARA and SDFS assessment were higher in patients with SCA27B than in those with ILOCA (75 vs 68.5 years and 75 vs 68 years, respectively; p-FDR = 0.001 and 0.003, respectively). Moreover, individual SARA item scores did not differ significantly between SCA27B and ILOCA after FDR correction (all p-FDR ≥ 0.091) (Supplementary Figure 3). An episodic disease course was more frequent in SCA27B (45.2% vs 12.7%, p-FDR < 0.001). Of note, episode features and triggers were similar among those patients with SCA27B and ILOCA with episodic ataxia. Core cerebellar manifestations (gait ataxia, dysarthria, and limb dysmetria) were similarly distributed between groups (all p-FDR ≥ 0.28). However, oculomotor abnormalities were more common in SCA27B than in ILOCA (82.5% vs 71.6%, p-FDR = 0.037), primarily owing to a higher frequency of downbeat nystagmus (67.4% vs 32.9%, p-FDR < 0.001). In contrast, ILOCA was associated with a greater burden of extracerebellar features, including spasticity, impaired light touch and pain sensation, reduced vibration sense, impaired proprioception, and hyperreflexia (all p-FDR ≤ 0.025). Other extracerebellar manifestations, including parkinsonism, tremor, dysautonomia, vestibular, cognitive, and other neurological features, did not differ significantly between groups (all p-FDR ≥ 0.125) (Table 2).

**Table 1.** Basal and demographic characteristics.

| **Table 1. Basal and demographic characteristics** |  |  |  |  |
| --- | --- | --- | --- | --- |
| **Variable** | **SCA27B**<br>**n=217** | **ILOCA**<br>**n=283** | **p** | **p-FDR** |
| Women, n/N (%) | 96/217 (44.2%) | 141/283 (49.8%) | 0.240 | 0.481 |
| Age at onset, median [Q1-Q3]/N | **63.00 (52.25–68.75) / 210** | **55.00 (46.00–64.00) / 266** | **<0.001** | **<0.001** |
| Age at evaluation, median [Q1-Q3]/N | **74.00 (65.00–80.00) / 207** | **66.00 (58.00–75.00) / 261** | **<0.001** | **<0.001** |
| Disease duration at evaluation, median [Q1-Q3]/N | 10.00 (6.00–15.00) / 200 | 9.00 (5.00–15.00) / 244 | 0.100 | 0.268 |
| SARA score, median [Q1-Q3]/N | 8.50 (6.00–14.00) / 155 | 10.00 (6.00–16.50) / 183 | 0.182 | 0.416 |
| Age at SARA, median [Q1-Q3]/N | **75.00 (69.00–80.00) / 117** | **68.50 (59.00–78.25) / 104** | **<0.001** | **0.001** |
| Disease duration at SARA, median [Q1-Q3]/N | 11.00 (8.00–16.00) / 113 | 11.00 (5.00–18.00) / 100 | 0.614 | 0.756 |
| SARA rate (SARA score/disease duration at evaluation),<br>median [Q1-Q3]/N | 0.81 (0.50–1.38) / 113 | 1.00 (0.42–1.54) / 99 | 0.551 | 0.734 |
| SDFS score, median [Q1-Q3]/N | 4.00 (3.00–5.00) / 158 | 3.00 (3.00–5.00) / 228 | 0.546 | 0.734 |
| Age at SDFS, median [Q1-Q3]/N | **75.00 (66.00–81.00) / 111** | **68.00 (59.00–78.00) / 141** | **<0.001** | **0.003** |
| Disease duration at SDFS, median [Q1-Q3]/N | 12.00 (7.00–16.00) / 109 | 10.50 (6.00–16.00) / 134 | 0.724 | 0.783 |
| SDFS rate (SDFS score/disease duration at | 0.33 (0.25–0.50) / 107 | 0.33 (0.20–0.57) / 133 | 0.734 | 0.783 |

| Table 1. Basal and demographic characteristics |  |  |  |  |
| --- | --- | --- | --- | --- |
| Variable | SCA27B<br>n=217 | ILOCA<br>n=283 | p | p-FDR |
| evaluation), median [Q1-Q3]/N |  |  |  |  |
| Ataxia disability score, median [Q1-Q3]/N | 2.00 (1.00–2.00) / 162 | 2.00 (1.00–2.00) / 232 | 0.800 | 0.800 |
| Age at Ataxia Disability Score, median [Q1-Q3]/N | **76.00 (66.00–81.00) / 97** | **68.50 (59.25–78.00) / 142** | **<0.001** | **0.001** |
| Disease duration at Ataxia Disability Score, median [Q1-Q3]/N | 12.00 (8.00–16.00) / 95 | 10.00 (6.00–16.00) / 135 | 0.309 | 0.549 |
| score/disease duration at evaluation, median [Q1-Q3]/N | 0.14 (0.10–0.22) / 93 | 0.15 (0.09–0.29) / 135 | 0.470 | 0.734 |

**Table 2.** Clinical features.

| **Table 2. Clinical features** |  |  |  |  |
| --- | --- | --- | --- | --- |
| **Parameter** | **SCA27B, n/N (%)**<br>**n=217** | **ILOCA, n/N (%)**<br>**n=283** | **p-value** | **p-FDR** |
| Episodic course | **98/217 (45.2%)** | **36/283 (12.7%)** | **<0.001** | **<0.001** |
| Trigger for the episodes |  |  |  |  |
| No trigger identified | 59/98 (60.2%) | 22/36 (61.1%) | 1.000 | 1.000 |
| Alcohol | 10/98 (10.2%) | 2/36 (5.6%) | 0.513 | 0.660 |
| Strenuous exercise | 20/98 (20.4%) | 5/36 (13.9%) | 0.462 | 0.660 |
| Other type of trigger | 21/98 (21.4%) | 11/36 (30.6%) | 0.360 | 0.649 |
| Type of episode |  |  |  |  |
| Gait impairment | 80/98 (81.6%) | 29/36 (80.6%) | 1.000 | 1.000 |
| Speech impairment | 54/98 (55.1%) | 16/36 (44.4%) | 0.331 | 0.649 |
| Visual disturbance | 31/98 (31.6%) | 6/36 (16.7%) | 0.126 | 0.567 |
| Vertigo | 19/98 (19.4%) | 11/36 (30.6%) | 0.241 | 0.649 |
| Gait ataxia | 180/192 (93.8%) | 266/280 (95.0%) | 0.682 | 0.855 |
| Impaired postural stability | 69/177 (39.0%) | 87/229 (38.0%) | 0.838 | 0.934 |
| Impaired tandem gait | 169/188 (89.9%) | 253/274 (92.3%) | 0.401 | 0.580 |
| Dysarthria | 113/190 (59.5%) | 176/280 (62.9%) | 0.499 | 0.671 |
| Dysphagia | 34/183 (18.6%) | 61/266 (22.9%) | 0.291 | 0.494 |
| **Parameter** | **SCA27B, n/N (%)**<br>**n=217** | **ILOCA, n/N (%)**<br>**n=283** | **p-value** | **p-FDR** |
| Oculomotor abnormalities | **170/206 (82.5%)** | **197/275 (71.6%)** | **0.007** | **0.037** |
| Downbeat nystagmus | **91/135 (67.4%)** | **49/149 (32.9%)** | **<0.001** | **<0.001** |
| Horizontal gaze-evoked nystagmus | 95/170 (55.9%) | 108/197 (54.8%) | 0.916 | 0.993 |
| Vertical gaze-evoked nystagmus | 33/170 (19.4%) | 26/197 (13.2%) | 0.118 | 0.282 |
| Paresis of horizontal gaze | 5/170 (2.9%) | 15/197 (7.6%) | 0.064 | 0.209 |
| Paresis of vertical gaze | 11/170 (6.5%) | 19/197 (9.6%) | 0.340 | 0.520 |
| Saccadic pursuit | 51/170 (30.0%) | 73/197 (37.1%) | 0.184 | 0.359 |
| Hypermetric saccades | 13/170 (7.6%) | 24/197 (12.2%) | 0.167 | 0.343 |
| Hypometric saccades | 21/170 (12.4%) | 37/197 (18.8%) | 0.114 | 0.282 |
| Slow saccades | 10/170 (5.9%) | 25/197 (12.7%) | 0.032 | 0.125 |
| Square-wave jerks | 3/170 (1.8%) | 4/197 (2.0%) | 1.000 | 1.000 |
| Finger-to-nose dysmetria | 123/192 (64.1%) | 189/279 (67.7%) | 0.428 | 0.597 |
| Heel-to-shin dysmetria | 129/190 (67.9%) | 208/278 (74.8%) | 0.116 | 0.282 |
| Parkinsonism | 16/187 (8.6%) | 30/261 (11.5%) | 0.347 | 0.520 |
| Tremor | 40/189 (21.2%) | 73/262 (27.9%) | 0.123 | 0.282 |
| Spasticity | **9/190 (4.7%)** | **50/260 (19.2%)** | **<0.001** | **<0.001** |
| **Parameter** | **SCA27B, n/N (%)**<br>**n=217** | **ILOCA, n/N (%)**<br>**n=283** | **p-value** | **p-FDR** |
| Impaired light touch and pain sensation | **4/170 (2.4%)** | **27/219 (12.3%)** | **<0.001** | **0.003** |
| Reduced vibration sense | **55/186 (29.6%)** | **109/252 (43.3%)** | **0.004** | **0.025** |
| Impaired proprioception | **7/164 (4.3%)** | **28/210 (13.3%)** | **0.004** | **0.025** |
| Neuropathic pain | 0/183 (0.0%) | 9/243 (3.7%) | 0.012 | 0.058 |
| Restless legs syndrome | 3/179 (1.7%) | 5/228 (2.2%) | 1.000 | 1.000 |
| Hyporeflexia | 34/189 (18.0%) | 67/256 (26.2%) | 0.051 | 0.182 |
| Hyperreflexia | **47/190 (24.7%)** | **107/263 (40.7%)** | **<0.001** | **0.004** |
| Extensor plantar response | 8/185 (4.3%) | 26/258 (10.1%) | 0.029 | 0.125 |
| Weakness/atrophy | 7/189 (3.7%) | 21/261 (8.0%) | 0.075 | 0.224 |
| Cough | 10/186 (5.4%) | 21/252 (8.3%) | 0.262 | 0.465 |
| Dysautonomia | 26/182 (14.3%) | 50/252 (19.8%) | 0.159 | 0.343 |
| Orthostatic hypotension | 7/26 (26.9%) | 14/50 (28.0%) | 1.000 | 1.000 |
| Urinary dysfunction | 14/26 (53.8%) | 34/50 (68.0%) | 0.316 | 0.514 |
| Hearing loss | 20/185 (10.8%) | 37/248 (14.9%) | 0.251 | 0.465 |
| Headache | 11/173 (6.4%) | 13/233 (5.6%) | 0.832 | 0.934 |
| Epilepsy | 2/122 (1.6%) | 4/160 (2.5%) | 0.701 | 0.855 |

| Table 2. Clinical features |  |  |  |  |
| --- | --- | --- | --- | --- |
| Parameter | SCA27B, n/N (%)<br>n=217 | ILOCA, n/N (%)<br>n=283 | p-value | p-FDR |
| Vestibular areflexia |  |  | 0.813 | 0.934 |
| No vestibular areflexia | 92/124 (74.2%); | 0: 146/190 (76.8%); |  |  |
| Positive HIT on clinical examination | 21/124 (16.9%); | 1: 27/190 (14.2%); |  |  |
| video-HIT positive | 11/124 (8.9%) | 2: 17/190 (8.9%) |  |  |
| Cognitive impairment |  |  | 0.532 | 0.692 |
| No cognitive decline | 157/185 (84.9%); | 216/259 (83.4%); |  |  |
| MCI | 24/185 (13.0%); | 32/259 (12.4%); |  |  |
| Dementia | 4/185 (2.2%) | 11/259 (4.2%) |  |  |

Compared with patients with ILOCA, those with SCA27B showed a lower frequency of cerebellar atrophy, involving both the vermis (56.2% vs 73.8% p-FDR = 0.001) and the cerebellar hemispheres (43.9% vs 64.1%, p-FDR < 0.001). Brainstem atrophy was also less frequent in SCA27B (2.9% vs 11.1%, p-FDR = 0.011), whereas the frequencies of cerebral atrophy, leukoencephalopathy, brainstem signal abnormalities, and spinal imaging abnormalities did not differ significantly between the two groups (Table 3).

**Table 3.**
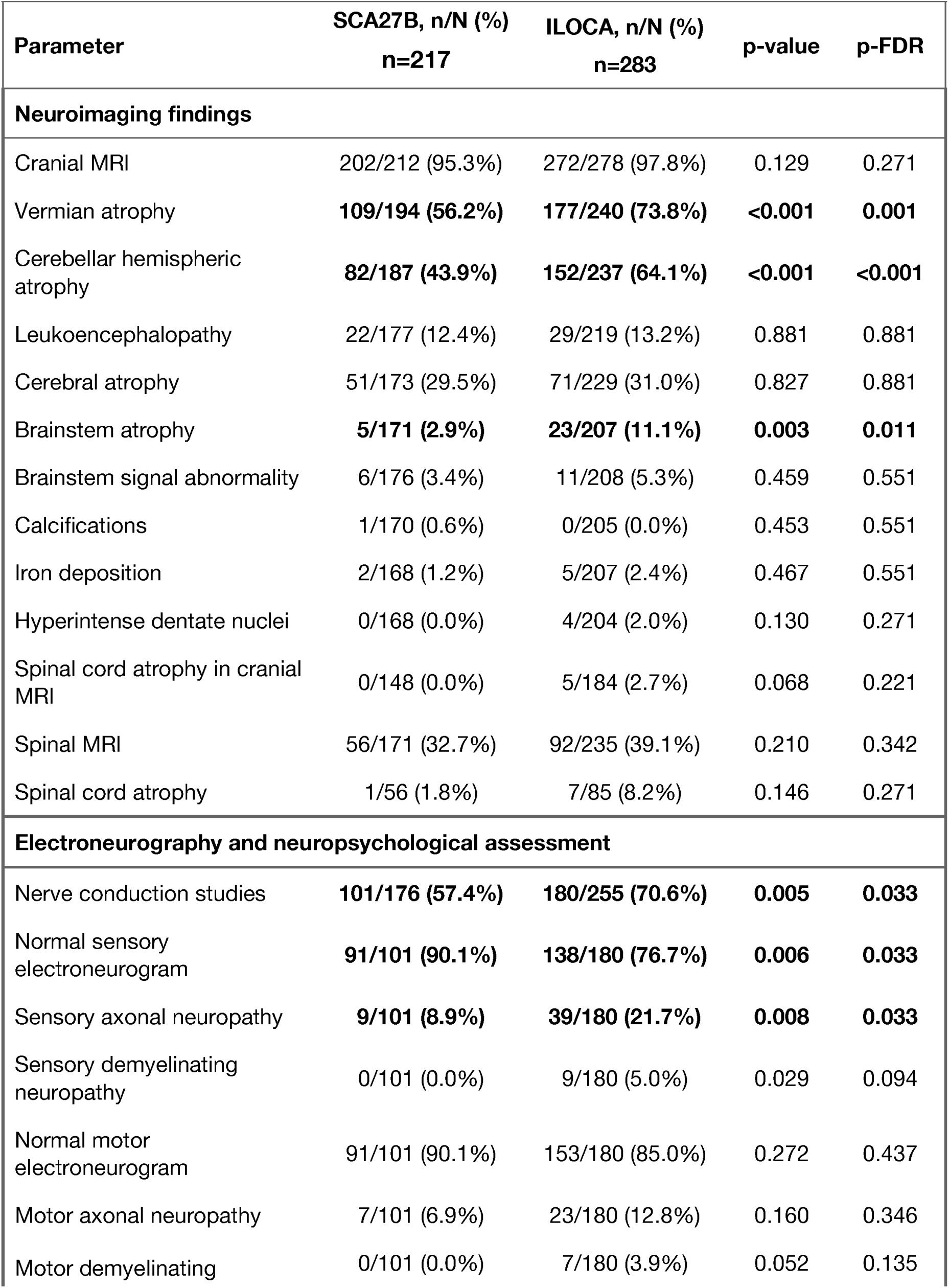
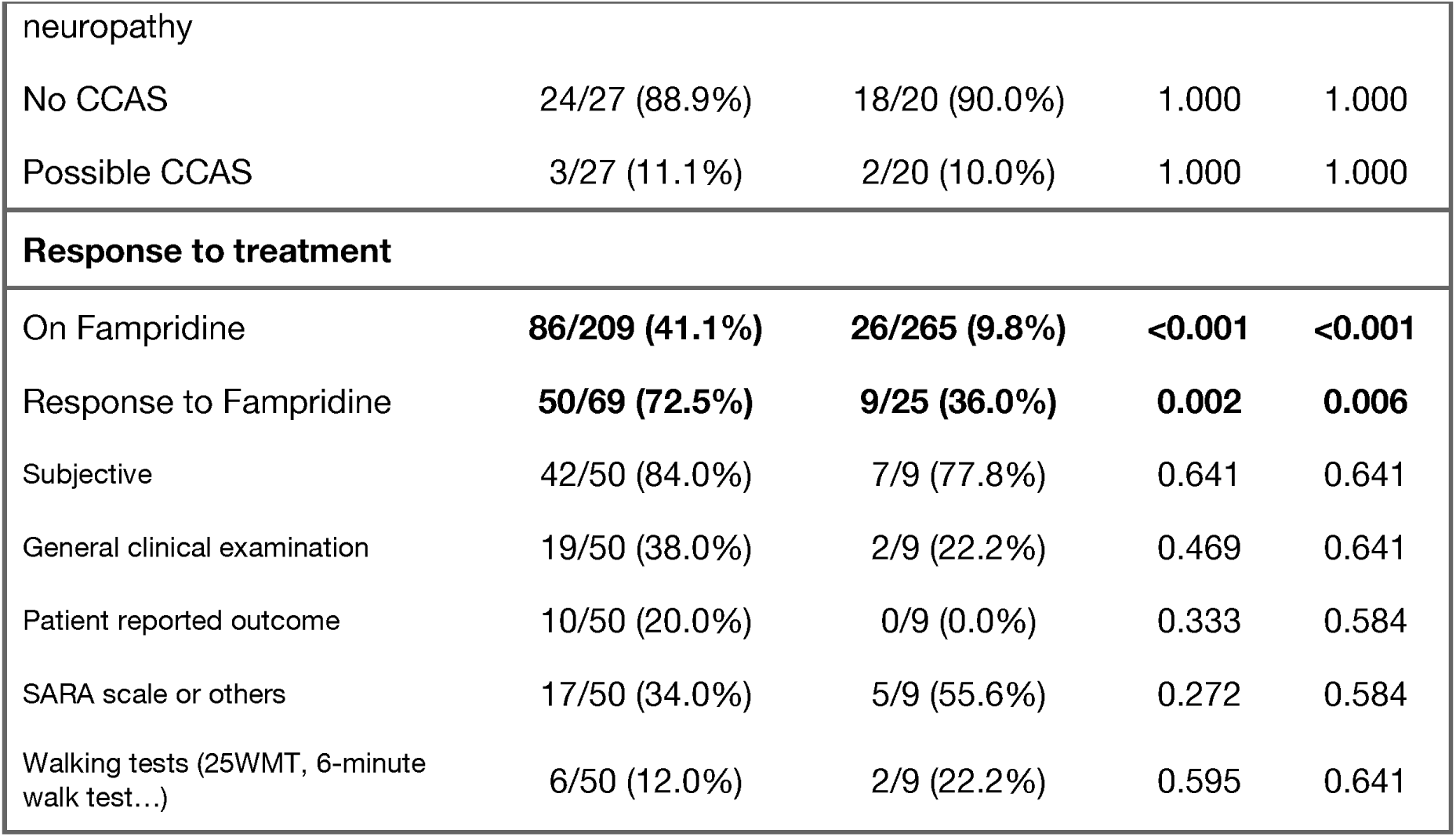
Neuroimaging findings, electroneurography, neuropsychological assessment, and response to treatment.

| Parameter | SCA27B, n/N (%)<br>n=217 | ILOCA, n/N (%)<br>n=283 | p-value | p-FDR |
| --- | --- | --- | --- | --- |
| **Neuroimaging findings** |  |  |  |  |
| Cranial MRI | 202/212 (95.3%) | 272/278 (97.8%) | 0.129 | 0.271 |
| Vermian atrophy | **109/194 (56.2%)** | **177/240 (73.8%)** | **<0.001** | **0.001** |
| Cerebellar hemispheric atrophy | **82/187 (43.9%)** | **152/237 (64.1%)** | **<0.001** | **<0.001** |
| Leukoencephalopathy | 22/177 (12.4%) | 29/219 (13.2%) | 0.881 | 0.881 |
| Cerebral atrophy | 51/173 (29.5%) | 71/229 (31.0%) | 0.827 | 0.881 |
| Brainstem atrophy | **5/171 (2.9%)** | **23/207 (11.1%)** | **0.003** | **0.011** |
| Brainstem signal abnormality | 6/176 (3.4%) | 11/208 (5.3%) | 0.459 | 0.551 |
| Calcifications | 1/170 (0.6%) | 0/205 (0.0%) | 0.453 | 0.551 |
| Iron deposition | 2/168 (1.2%) | 5/207 (2.4%) | 0.467 | 0.551 |
| Hyperintense dentate nuclei | 0/168 (0.0%) | 4/204 (2.0%) | 0.130 | 0.271 |
| Spinal cord atrophy in cranial MRI | 0/148 (0.0%) | 5/184 (2.7%) | 0.068 | 0.221 |
| Spinal MRI | 56/171 (32.7%) | 92/235 (39.1%) | 0.210 | 0.342 |
| Spinal cord atrophy | 1/56 (1.8%) | 7/85 (8.2%) | 0.146 | 0.271 |
| **Electroneurography and neuropsychological assessment** |  |  |  |  |
| Nerve conduction studies | **101/176 (57.4%)** | **180/255 (70.6%)** | **0.005** | **0.033** |
| Normal sensory electroneurogram | **91/101 (90.1%)** | **138/180 (76.7%)** | **0.006** | **0.033** |
| Sensory axonal neuropathy | **9/101 (8.9%)** | **39/180 (21.7%)** | **0.008** | **0.033** |
| Sensory demyelinating neuropathy | 0/101 (0.0%) | 9/180 (5.0%) | 0.029 | 0.094 |
| Normal motor electroneurogram | 91/101 (90.1%) | 153/180 (85.0%) | 0.272 | 0.437 |
| Motor axonal neuropathy | 7/101 (6.9%) | 23/180 (12.8%) | 0.160 | 0.346 |
| Motor demyelinating | 0/101 (0.0%) | 7/180 (3.9%) | 0.052 | 0.135 |

| **Parameter** | **SCA27B, n/N (%)**<br>**n=217** | **ILOCA, n/N (%)**<br>**n=283** | **p-value** | **p-FDR** |
| --- | --- | --- | --- | --- |
| neuropathy |  |  |  |  |
| No CCAS | 24/27 (88.9%) | 18/20 (90.0%) | 1.000 | 1.000 |
| Possible CCAS | 3/27 (11.1%) | 2/20 (10.0%) | 1.000 | 1.000 |
| **Response to treatment** |  |  |  |  |
| On Fampridine | **86/209 (41.1%)** | **26/265 (9.8%)** | **<0.001** | **<0.001** |
| Response to Fampridine | **50/69 (72.5%)** | **9/25 (36.0%)** | **0.002** | **0.006** |
| Subjective | 42/50 (84.0%) | 7/9 (77.8%) | 0.641 | 0.641 |
| General clinical examination | 19/50 (38.0%) | 2/9 (22.2%) | 0.469 | 0.641 |
| Patient reported outcome | 10/50 (20.0%) | 0/9 (0.0%) | 0.333 | 0.584 |
| SARA scale or others | 17/50 (34.0%) | 5/9 (55.6%) | 0.272 | 0.584 |
| Walking tests (25WMT, 6-minute walk test...) | 6/50 (12.0%) | 2/9 (22.2%) | 0.595 | 0.641 |

Peripheral nerve involvement was less frequent in patients with SCA27B than in those with ILOCA. Nerve conduction studies were performed in 101/176 (57.4%) patients with SCA27B and 180/255 (70.6%) with ILOCA. Among those who underwent testing, normal sensory electroneurography was more frequent in SCA27B than in ILOCA (90.1% vs 76.7%, p-FDR = 0.033). Sensory axonal neuropathy was more common in ILOCA (8.9% vs 21.7%, p-FDR = 0.033). Sensory demyelinating neuropathy was observed only in ILOCA (0% vs 5.0%), although this difference did not remain significant after FDR correction (p-FDR = 0.094). Motor nerve conduction findings were comparable between groups, including the frequency of normal motor electroneurography (90.1% vs 85.0%, p-FDR = 0.437). Cognitive-affective function was assessed using the Spanish version of the Cerebellar Cognitive Affective Syndrome Scale (CCAS Scale) in 47 patients (16,17).The distribution of CCAS categories was comparable between groups (p = 1.000, p-FDR = 1.000), with no CCAS in 24/27 (88.9%) patients with SCA27B and 18/20 (90.0%) with ILOCA (p-FRD = 1.000), and possible CCAS in 3/27 (11.1%) and 2/20 (10.0%), respectively (Table 3).

4-AP use was more frequent among patients with SCA27B than among those with ILOCA (41.1% vs. 9.8%, p-FDR < 0.001), reflecting its preferential use in SCA27B. Among treated patients, the proportion of responders was higher in SCA27B than in ILOCA (72.5% vs. 36.0%, p-FDR = 0.006). Most responses were based on subjective patient-reported improvement in both groups (84.0% in SCA27B and 77.8% in ILOCA, p-FDR = 0.641) (Table 3). Response of episodic symptoms to treatment was not systematically assessed.

### Clinical characterization of the Spanish SCA27B cohort

We compared clinical and imaging features among three groups of patients with SCA27B according to repeat size: 200–249 (n=17), 250–299 (n=51), and ≥300 GAA-TTC repeats (n=149). Overall, demographic characteristics, disease onset, disease duration, clinical severity and progression measures and clinical features were comparable across repeat-length groups (Table 4). Triplet repeat count was not associated with age at onset (Pearson’s r = −0.031, p = 0.655; linear regression R² = 0.001) (Figure 1B). When correcting for multiple comparisons, neuroimaging findings were comparable in patients with 200–249, 250–299, and >300 repeats (Table 4). No significant differences were observed in electroneurography findings either (all p-FDR ≥ 0.681).

**Table 4.**
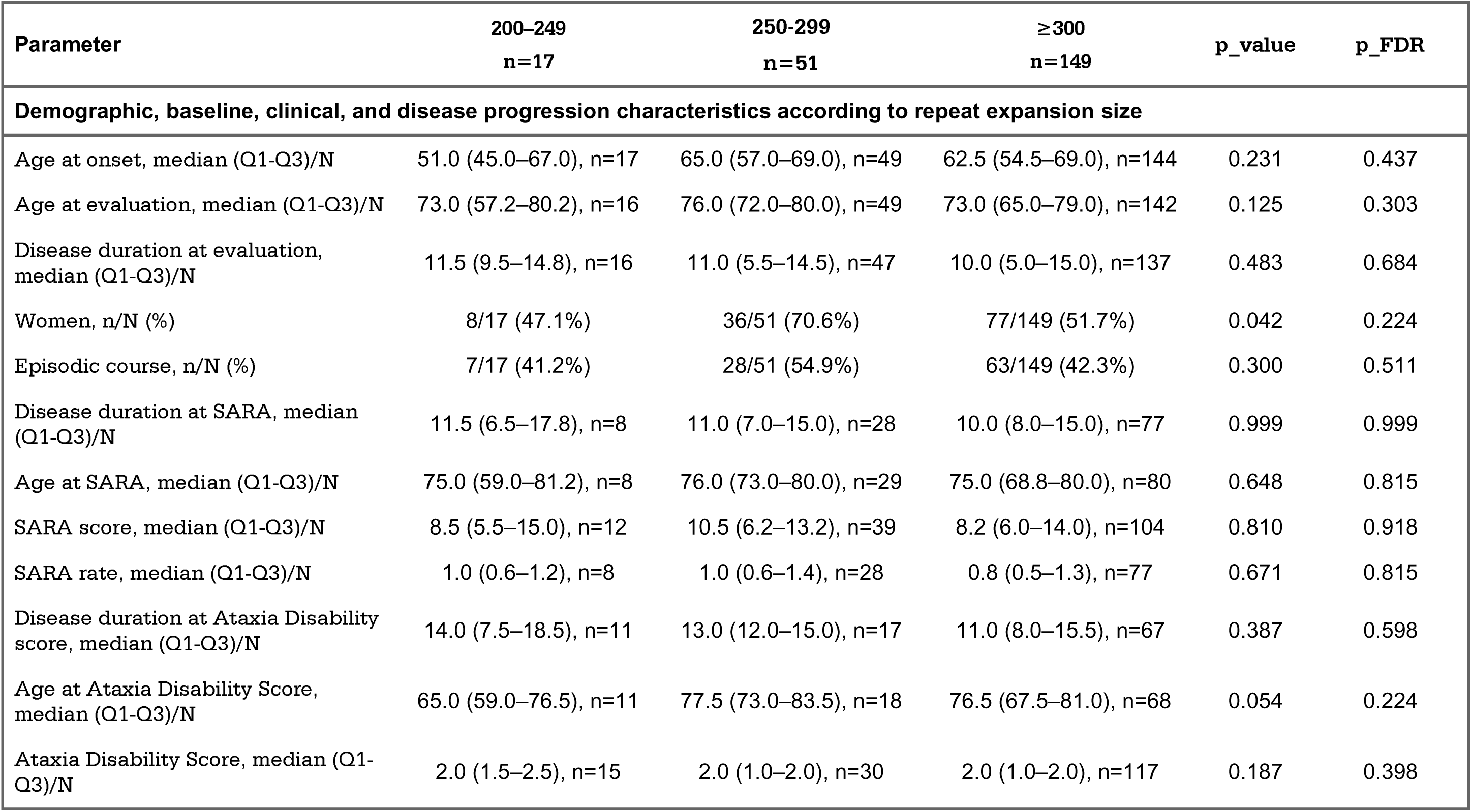
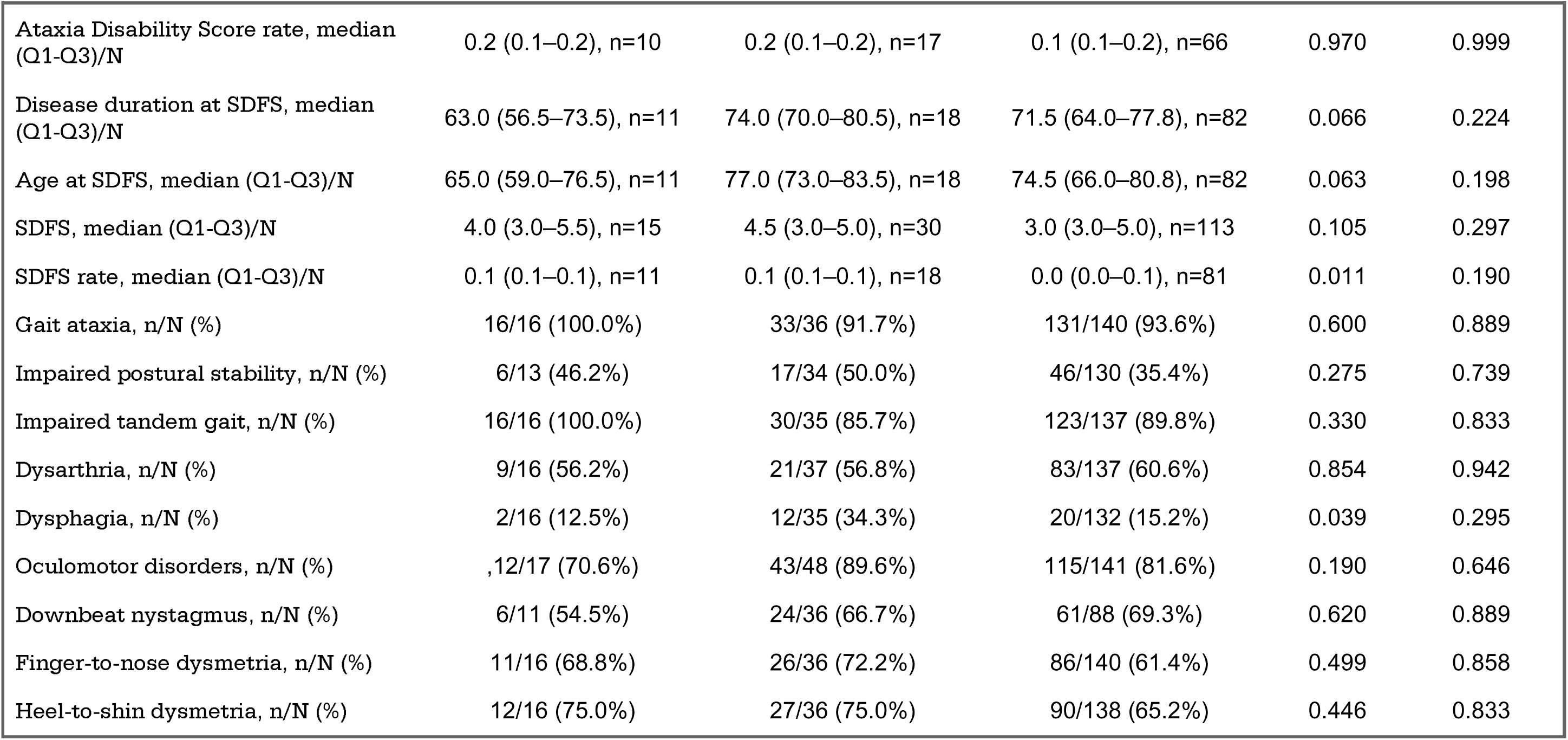
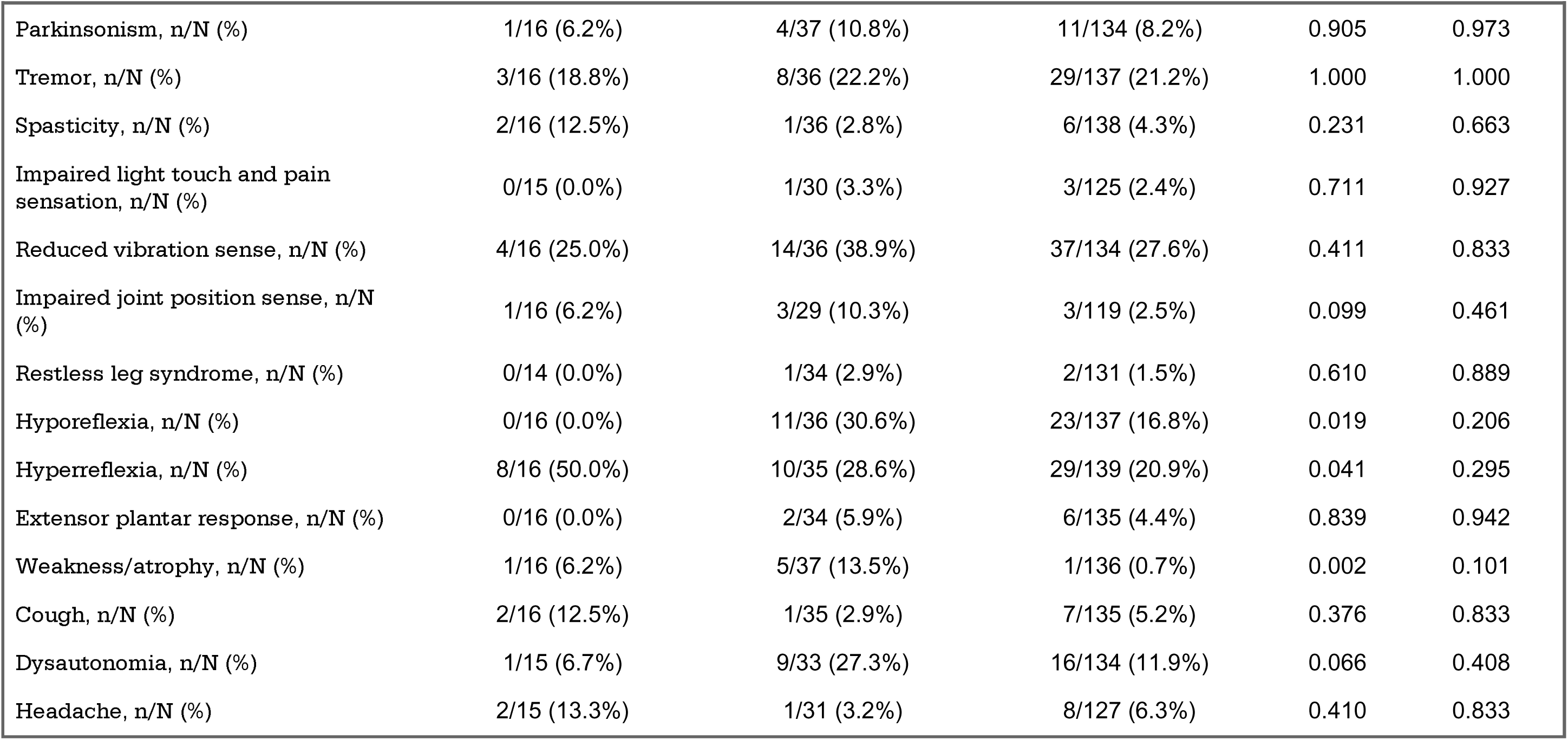
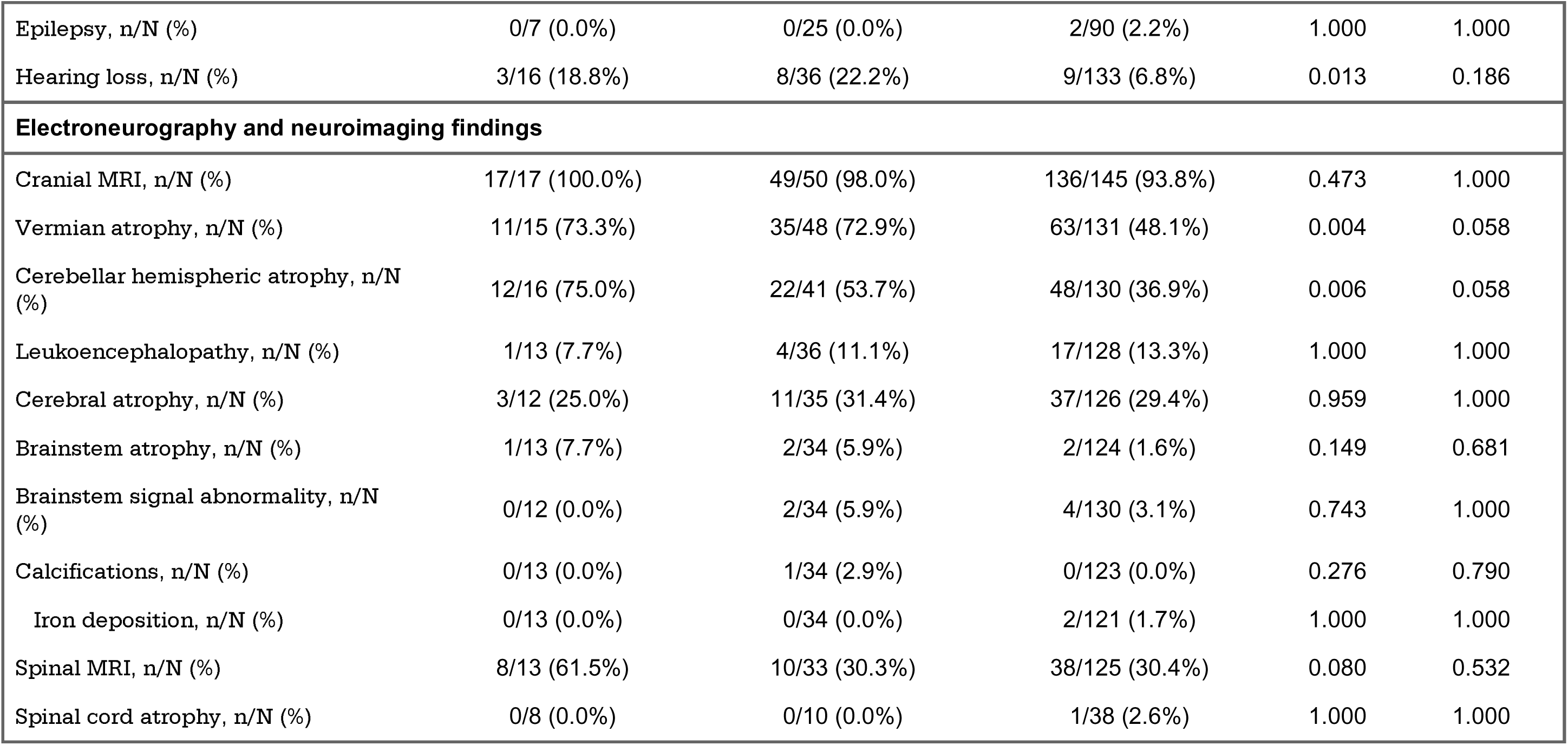
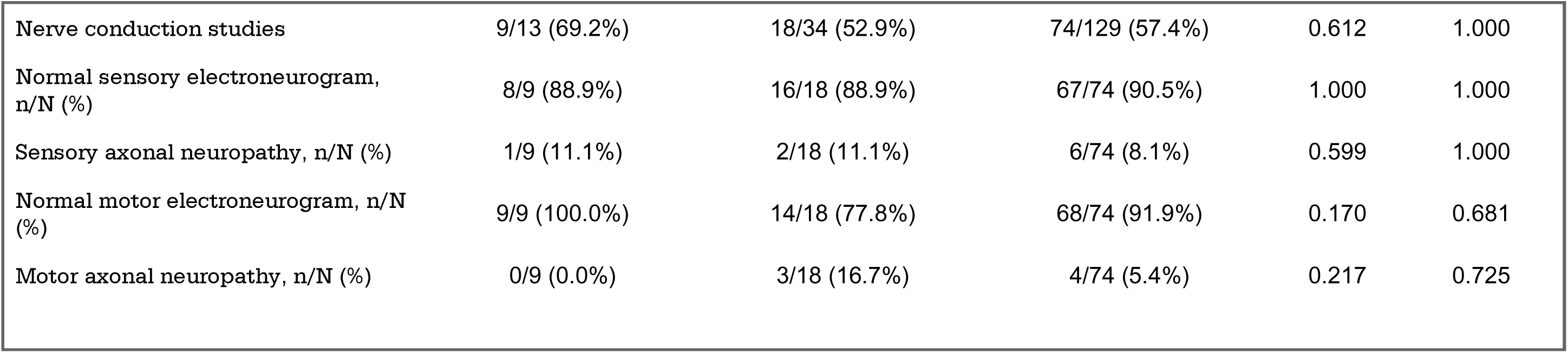
Demographic, clinical, disease progression, and ancillary assessment findings according to repeat expansion size.

From the 217 patients with SCA27B, 14 patients carried biallelic >200 GAA-TTC repeat expansions. Monoallelic (n=203) and biallelic (n=14) expansion carriers showed largely similar phenotypes (all p-FDR ≥ 0.809) (Supplementary Table 2). Neuroimaging and neurophysiological measures were also comparable between groups (all p-FDR = 1.000) (Supplementary Table 3).

Intergenerational instability was assessed in 12 parent–offspring transmissions involving 8 unique parents (3 females and 5 males). Instability was defined as the difference between offspring and parental repeat sizes, with positive values indicating expansions and negative values indicating contractions. All four maternal transmissions resulted in expansions, with a median instability of +17.5 GAA-TTC repeats (range, +8 to +33), whereas all eight paternal transmissions resulted in contractions, with a median instability of −133.5 GAA-TTC repeats (range, −182 to −23). The distribution of expansions and contractions differed significantly according to parental sex (Fisher’s exact test, p = 0.002). To account for the fact that some parents contributed more than one transmission, instability was additionally summarized at the unique-parent level by calculating the mean instability for each parent. Among the 3 female and 5 male parents, the median parent-specific mean instability was +19 GAA-TTC repeats (range, +8 to +24.5) for females and −126 GAA-TTC repeats (range, −167 to −25) for males. This difference was statistically significant (Wilcoxon rank-sum test, W = 15, p = 0.036). Finally, although a trend was observed, there was no significant association between parental expansion size and parent-specific mean intergenerational instability (Spearman’s ρ = −0.286, p = 0.501) (Supplementary Figure 4A and 4B).

### Longitudinal analyses

The longitudinal cohort comprised 190 patients contributing 372 SARA assessments, including 146 patients with SCA27B (258 assessments) and 44 with ILOCA (114 assessments). Patients with SCA27B had a significantly later age at onset (median 63.0 vs. 54.0 years, p = 0.032) and were older at baseline assessment (median 73.0 vs. 65.0 years, p = 0.028) than patients with ILOCA, whereas disease duration (median 9.0 vs. 9.0 years, p = 1.000) and the proportion of female patients (41.7% vs. 50.0%, p = 0.386) were similar between groups.

In a linear mixed-effects model, SARA scores increased significantly with disease duration (β = 0.397 points/year, 95% CI 0.251–0.543; p < 0.001), with, no evidence of a different rate of progression between SCA27B and ILOCA (disease duration-by-diagnosis interaction β = −0.024, 95% CI −0.203 to 0.155; p = 0.795; Figure 1C). The estimated annual increase in SARA score was 0.397 points/year (95% CI 0.251–0.543) for ILOCA and 0.373 points/year (95% CI 0.270–0.477) for SCA27B. After adjustment for age at assessment, disease duration remained significantly associated with higher SARA scores (β = 0.348 points/year, 95% CI 0.200–0.497; p < 0.001). The estimated annual increase was 0.348 points/year (95% CI 0.200–0.497) for ILOCA and 0.296 points/year (95% CI 0.182–0.411) for SCA27B, with no significant difference in progression rates between groups (disease duration-by-diagnosis interaction β = −0.052, 95% CI −0.229 to 0.125; p = 0.564). Age at assessment was independently associated with higher SARA scores (β = 0.116 points/year, 95% CI 0.040–0.192; p = 0.003).

Among patients with SCA27B, neither sex nor the sex-by-disease duration interaction was significantly associated with SARA progression in the full longitudinal cohort after adjustment for age at assessment (sex effect: β = 2.559, 95% CI −0.721 to 5.839; p = 0.128; sex-by-disease duration interaction: β = −0.203, 95% CI −0.437 to 0.031; p = 0.089) (Figure 1D). However, when the analysis was restricted to patients with two or more study visits, a significant sex-by-disease duration interaction was observed after adjustment for age at assessment (β = −0.554, 95% CI −0.863 to −0.246; p < 0.001). The estimated annual increase in SARA score was 0.710 points/year (95% CI 0.416–1.003; p < 0.001) in women and 0.155 points/year (95% CI −0.011 to 0.322; p = 0.067) in men, indicating a significantly slower rate of SARA progression in men than in women (Supplementary Figure 5).

We also analyzed the association between 4-AP treatment and disease progression as measured by the SARA score. A total of 250 longitudinal SARA assessments from 134 patients with SCA27B were included, comprising 155 evaluations in patients not receiving 4-AP and 95 in patients receiving 4-AP. Twenty patients changed treatment status during follow-up. In a linear mixed-effects model adjusted for age at evaluation, longer disease duration was significantly associated with higher SARA scores (β = 0.25 points/year, 95% CI 0.11–0.39; p < 0.001). In contrast, neither 4-AP treatment (β = 0.27, 95% CI −1.91 to 2.44; p = 0.809) nor its interaction with disease duration (β = −0.01, 95% CI −0.19 to 0.17; p = 0.893) was significantly associated with SARA scores. Estimated SARA progression rates were 0.25 points/year (95% CI, 0.11–0.39) in patients not receiving 4-AP and 0.24 points/year (95% CI, 0.06–0.41) in patients receiving 4-AP. The difference between slopes was not significant (−0.01 points/year; 95% CI, −0.19 to 0.17; p = 0.893), indicating no detectable association between 4-AP treatment and the rate of SARA progression (Figure 1E).

To further explore the potential association between 4-AP treatment and disease progression, we performed a longitudinal within-patient analysis including 20 patients with SCA27B who changed treatment status during follow-up, contributing 73 SARA assessments. In the mixed-effects model adjusted for age at evaluation, longer disease duration was significantly associated with increasing SARA scores (β = 0.42 points/year, 95% CI 0.06–0.78; p = 0.022). However, neither 4-AP treatment (β = −0.49, 95% CI −3.45 to 2.47; p = 0.741) nor the interaction between treatment and disease duration (β = −0.03, 95% CI −0.29 to 0.24; p = 0.853) was significantly associated with SARA scores. Estimated SARA progression rates were 0.42 points/year (95% CI, 0.06–0.78) in patients not receiving 4-AP and 0.40 points/year (95% CI, 0.06–0.73) in patients receiving 4-AP. The difference between slopes was not significant (−0.03 points/year; 95% CI, −0.29 to 0.24; p = 0.853), providing no evidence of an association between 4-AP treatment and disease progression (Figure 1F).

## Discussion

The identification of FGF14 GAA-TTC repeat expansions has substantially reshaped the diagnostic landscape of LOCA (1,2). In this nationwide multicenter study, pathogenic FGF14 expansions were identified in 25.8% of patients with unexplained LOCA, and our cohort of 217 genetically confirmed patients with SCA27B represents one of the largest cohorts reported to date. Direct comparison with a large ILOCA cohort allowed us to define clinical features distinguishing SCA27B from idiopathic disease and, together with the longitudinal analyses, provided further insight into disease progression, repeat-size effects, intergenerational instability, and treatment response.

The high diagnostic yield reinforces the contribution of *FGF14* expansions to unexplained LOCA in Spain and supports SCA27B as one of the most frequent inherited causes of LOCA in individuals of European ancestry (18–20). Across cohorts, the reported frequency of *FGF14* GAA-TTC repeat expansions in patients with unexplained LOCA has varied considerably, ranging from approximately 5% to over 30%, while the highest frequencies have been reported in French Canadian patients (45–65%) (1,3,21). The 26% diagnostic yield observed in our nationwide Spanish cohort is therefore within the range reported in other European populations and is consistent with findings from smaller previous Spanish cohorts (3,22).

In Spain, a nationwide epidemiological survey of 1,371 patients with hereditary ataxia performed before the discovery of *RFC1* and *FGF14* expansions reported that almost half of patients remained without a molecular diagnosis, reflecting the limitations of diagnostic algorithms available at that time (23). The most common causes identified were Friedreich ataxia (249/1,371, 18.2%) and spinocerebellar ataxia type 3 (SCA3; 131/1,371, 9.6%). By comparison, *FGF14* repeat expansions accounted for 111 of 430 patients (25.8%) in our LOCA cohort. Although these cohorts are not directly comparable because our study specifically recruited patients with late-onset cerebellar ataxia, the high diagnostic yield and the identification of 217 genetically confirmed patients with SCA27B highlight its substantial contribution to the genetic landscape of LOCA in Spain. These findings illustrate how the recognition of recently identified repeat expansion disorders is reshaping the molecular diagnosis of adult-onset cerebellar ataxia and support the incorporation of *FGF14* repeat expansion testing into diagnostic algorithms for unexplained LOCA (1,2,24,25).

Our findings also provide a detailed characterization of the phenotype distinguishing SCA27B from ILOCA. SCA27B was associated with later disease onset (median 63 years of age), more frequent episodic manifestations, and a higher frequency of downbeat nystagmus, whereas extracerebellar manifestations, particularly pyramidal and sensory abnormalities, were more frequent in ILOCA. This agrees with previous studies comparing SCA27B and ILOCA based on substantially smaller numbers of patients (10,22,26) and provide useful clinical clues for prioritizing *FGF14* testing in clinical practice.

The neuroimaging and neurophysiological findings further support this predominantly cerebellar phenotype. Cerebellar and brainstem atrophy and sensory neuropathy were less frequent in SCA27B than in ILOCA, despite similar disease duration and clinical severity. These findings are consistent with previous reports describing relatively preserved cerebellar structure in many patients with SCA27B (1,2,24). This is completely different from other SCAs where cerebellar atrophy may even precede clinical symptoms (27,28). The absence of marked cerebellar atrophy should therefore not argue against the diagnosis in a patient with a compatible clinical phenotype. Although our sample was small, CCAS was uncommon and did not differ between groups in our cohort. However, a recent study identified a significant proportion of cognitive-affective abnormalities in a series of 17 patients with SCA27B (29), highlighting the need for additional studies in larger cohorts.

The association between repeat size and severity remains less clear. In contrast to many repeat expansion disorders (30–32), we found no association between repeat length and age at onset, overall disease severity, or most clinical measures (1,2,24). However, this relationship remains controversial, as some groups have reported an inverse correlation between GAA-TTC repeat length and age at onset (1,2,33). Patients carrying 200–249 repeats showed broadly similar clinical characteristics to those with larger expansions. Intermediate alleles should therefore be interpreted in conjunction with the clinical phenotype and other available genetic and familial information. Similarly, biallelic carriers in our cohort did not show significant phenotypic differences from monoallelic carriers after correction for multiple comparisons, despite the theoretical expectation of a gene dosage effect, although the small number of biallelic patients limits definitive conclusions (7).

Intergenerational repeat instability has been recognized previously (34), with maternal transmissions tending to result in expansion and paternal transmissions in contraction. In our cohort, we further confirmed this sex-dependent pattern. The biological basis of this parent-of-origin effect remains uncertain but may involve sex-specific mechanisms of germline repeat stability, DNA repair, or meiotic recombination. Although the small number of informative transmissions precludes firm conclusions regarding individual transmission risk, these findings support consideration of parental sex when interpreting *FGF14* repeat transmission.

Our longitudinal analysis of SARA provided further evidence that SCA27B is a slowly progressive disorder. SARA scores increased with disease duration in both SCA27B and ILOCA, with estimated progression rates of 0.296 and 0.348 points/year, respectively, after adjustment for age at assessment, with no significant difference between groups. This estimate is consistent with previous studies, which reported annual SARA progression rates between 0.23 and 0.54 points/year (7,9,10,35).

Sex was not associated with SARA progression in the overall SCA27B cohort. However, when only patients with at least two study visits were analysed, a significant sex-by-disease-duration interaction indicated slower progression in men than women. This finding has also been suggested by other authors (36) but not confirmed in larger studies (6,35). Therefore, it should be interpreted cautiously, especially because the restricted analysis involving a smaller and selected subgroup in this study.

4-AP was more frequently used in SCA27B and a greater proportion of treated patients were classified as responders than in ILOCA. However, neither treatment nor its interaction with disease duration was associated with lower SARA scores in the longitudinal analyses. The within-patient analysis of individuals who changed treatment status during follow-up yielded similar results, with comparable SARA progression rates during treated and untreated periods. However, these findings should not be interpreted as evidence against symptomatic benefit. SARA may be relatively insensitive to transient or domain-specific improvements, particularly in gait, oscillopsia, or episodic symptoms. The observational design and lack of systematic assessment of episodic manifestations further limit objective response assessment. Future studies should therefore incorporate treatment-sensitive clinical, patient-reported, and digital outcome measures.

The strengths of this study include its nationwide multicenter design, large genetically confirmed SCA27B cohort, substantial ILOCA comparator group, and integration of clinical, imaging, neurophysiological, intergenerational, and longitudinal data. Several limitations should nevertheless be considered. Part of the clinical data was collected retrospectively, ancillary investigations were not uniformly available across centers, and MRI protocols were not standardized. Moreover, the degree of cerebellar atrophy was assessed qualitatively rather than quantitatively, without standardized volumetric or other objective measures. In particular, potentially informative imaging signs such as the superior cerebellar peduncle sign (37) were not systematically assessed.

In conclusion, FGF14 repeat expansions accounted for approximately one quarter of previously unexplained LOCA in this nationwide Spanish cohort, supporting SCA27B as one of the most frequent genetic causes of LOCA in Spain. SCA27B showed a distinctive clinical profile characterized by later onset (median 63 years of age), episodic manifestations, downbeat nystagmus, limited extracerebellar involvement, and relatively preserved structural imaging. Repeat size was not clearly associated with age at onset or disease severity, whereas intergenerational transmission showed a parent-of-origin effect. Longitudinally, SARA scores showed that SCA27B followed a slowly progressive course comparable to ILOCA. 4-AP was not able to modify the SARA course in our study despite the high rate of responders based on patient-report and general exam assessments. These findings provide clinically relevant benchmarks for future studies of phenotypic delineation, progression, and treatment response.

## Acknowledgements

Authors want to acknowledge all the patients and families involved in this study and the collaboration of the different biobanks participating in this project, including the Basque Biobank -BIOEF; and the ISABIAL biobank. We thank CERCA Programme/Generalitat de Catalunya for institutional support, the IMPaCT Genómica 2 project, the Secretariat for Universities and Research of the Ministry of Business and Knowledge of the government of Catalonia (2021SGR00899), and the Association ASL-HSP to A.P.; and the Instituto de Salud Carlos III (ISCIII) and “Fondo Europeo de Desarrollo Regional (FEDER), Unión Europea, una manera de hacer Europa” (FIS PI23/01090) to C.C. The ISCIII also supported V.V.-S. (Rio Hortega, CM18/ 00145). Additional funding from the “La Marató de TV3” Foundation (202006-30) to C.C. and A.P. was obtained. Pujol, Casasnovas, Velez-Santamaria and Fourcade belong to the REMMA program for Malalties Minoritaries de l’Adult (Rare Diseases) of IDIBELL. We also appreciate the support of the Spanish Society of Neurology providing access and support to the RedCap database used in this project. A. V. has a grant from the Gipuzkoa College of Physicians. D.C.-C. has a grant from III Carlos Health Institute (Río Hortega, CM24/00220). D.P. holds a Fellowship Award from the Canadian Institutes of Health Research.

## Data sharing

The data that support the findings of this study are available from the corresponding author upon reasonable request. Individual participant data are not publicly available due to ethical and legal restrictions related to participant confidentiality.

## Figure Legends

**Supplementary Figure 1** (A) Geographic distribution of participating centers across Spain. (B) Study inclusion flowchart. Among 430 patients with unexplained late-onset cerebellar ataxia (LOCA) tested for FGF14 GAA–TTC repeat expansions, 90 additional genetically confirmed Spanish SCA27B patients from external laboratories were included.

**Supplementary Figure 2.** Patients with SCA27B showed a significant later age at onset compared to patients with ILOCA (median 63 vs 55 years, p < 0.001).

**Supplementary Figure 3. Effect sizes for individual SARA items between ILOCA and SCA27B.** Cliff’s delta (δ) with 95% bootstrap confidence intervals is shown. Positive values indicate higher ILOCA scores, negative values higher SCA27B scores, and the dashed line no effect (δ = 0).

**Supplementary Figure 4. Sex-specific intergenerational transmission of the SCA27B repeat expansion.** (A) Intergenerational instability of the FGF14 GAA repeat expansion by parental sex. Repeat-length change was calculated as offspring minus parental repeat size. (B) Relationship between parental allele size and repeat-length change. Positive values indicate expansions; negative values contractions. Dashed line indicates no change.

**Supplementary Figure 5.** Longitudinal SARA trajectories by disease duration in women and men with SCA27B with at least two assessments. Points represent individual observations, thin lines individual trajectories, and thick lines with shaded areas mixed-effects model predictions and 95% confidence intervals.

