## Supplementary figures and images for "A Large Spanish Cohort Study Defines SCA27B Distinct Clinical Phenotype and its Longitudinal Progression"

### Supplementary Figure 1

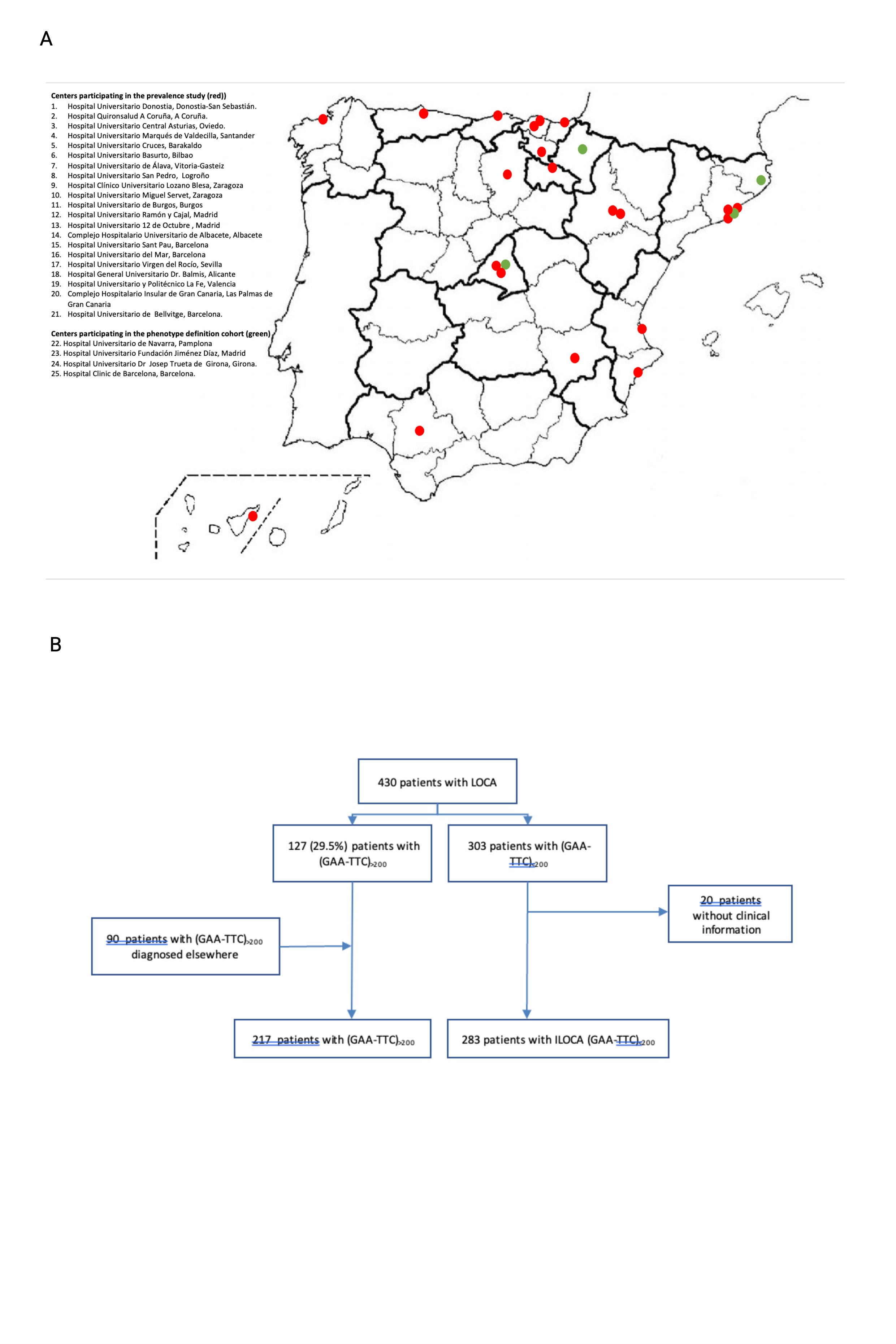

### Supplementary Figure 2

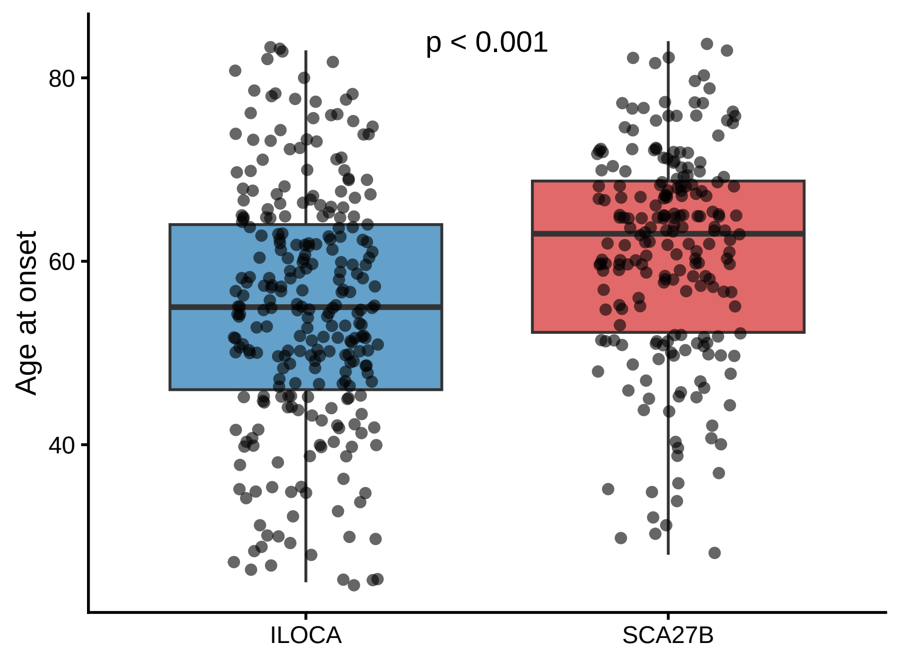

### Supplementary Figure 3

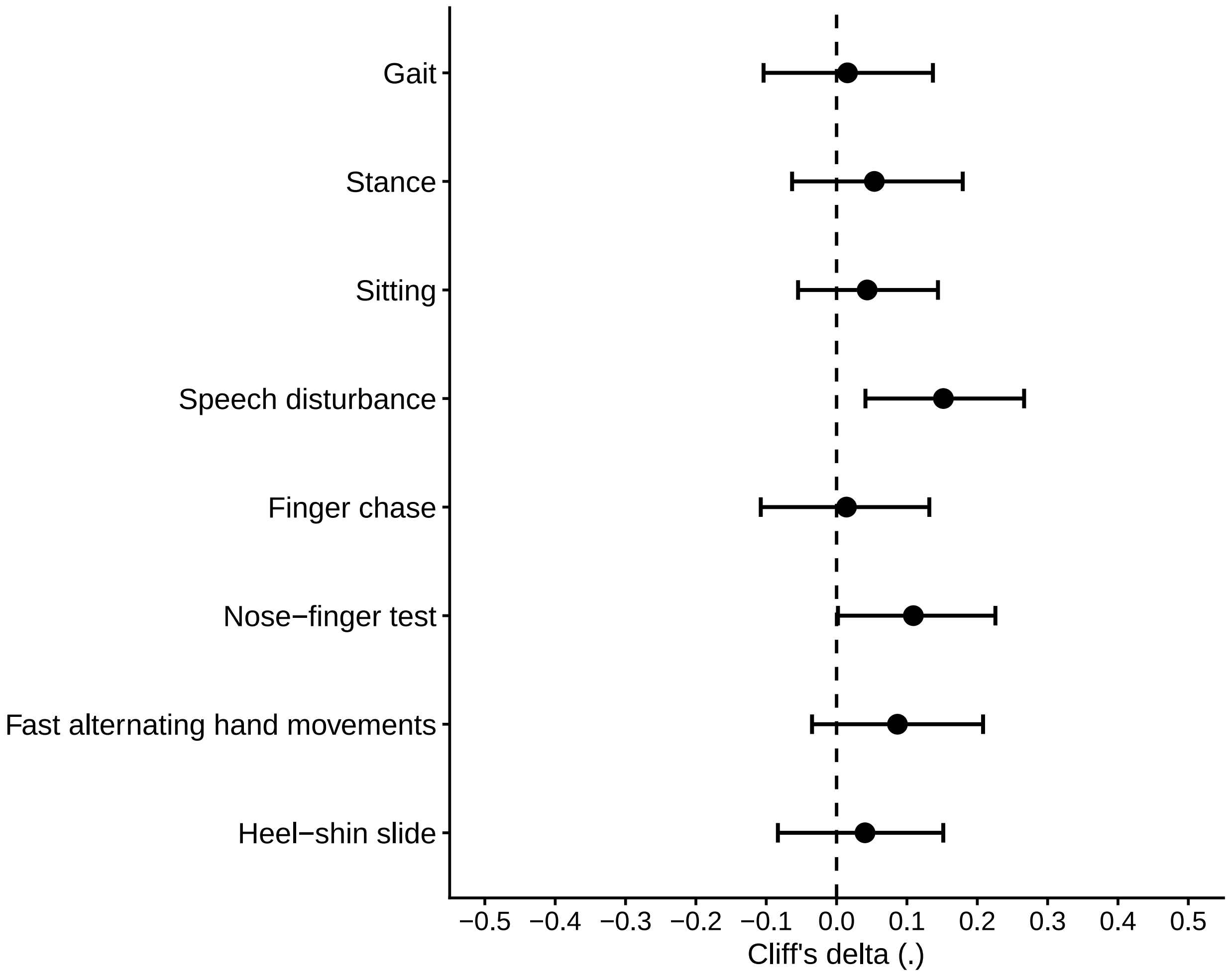

### Supplementary Figure 4

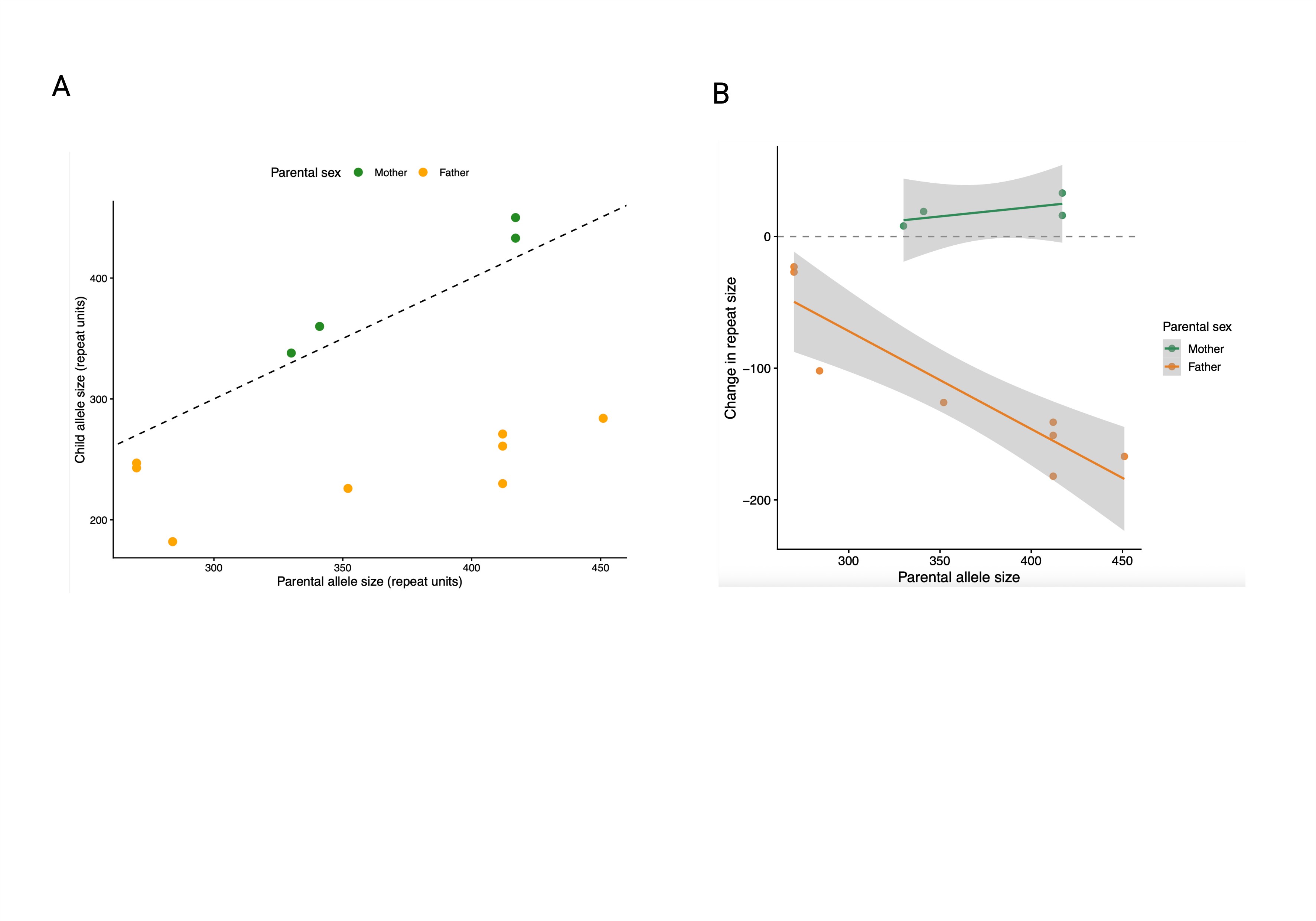

### Supplementary Figure 5

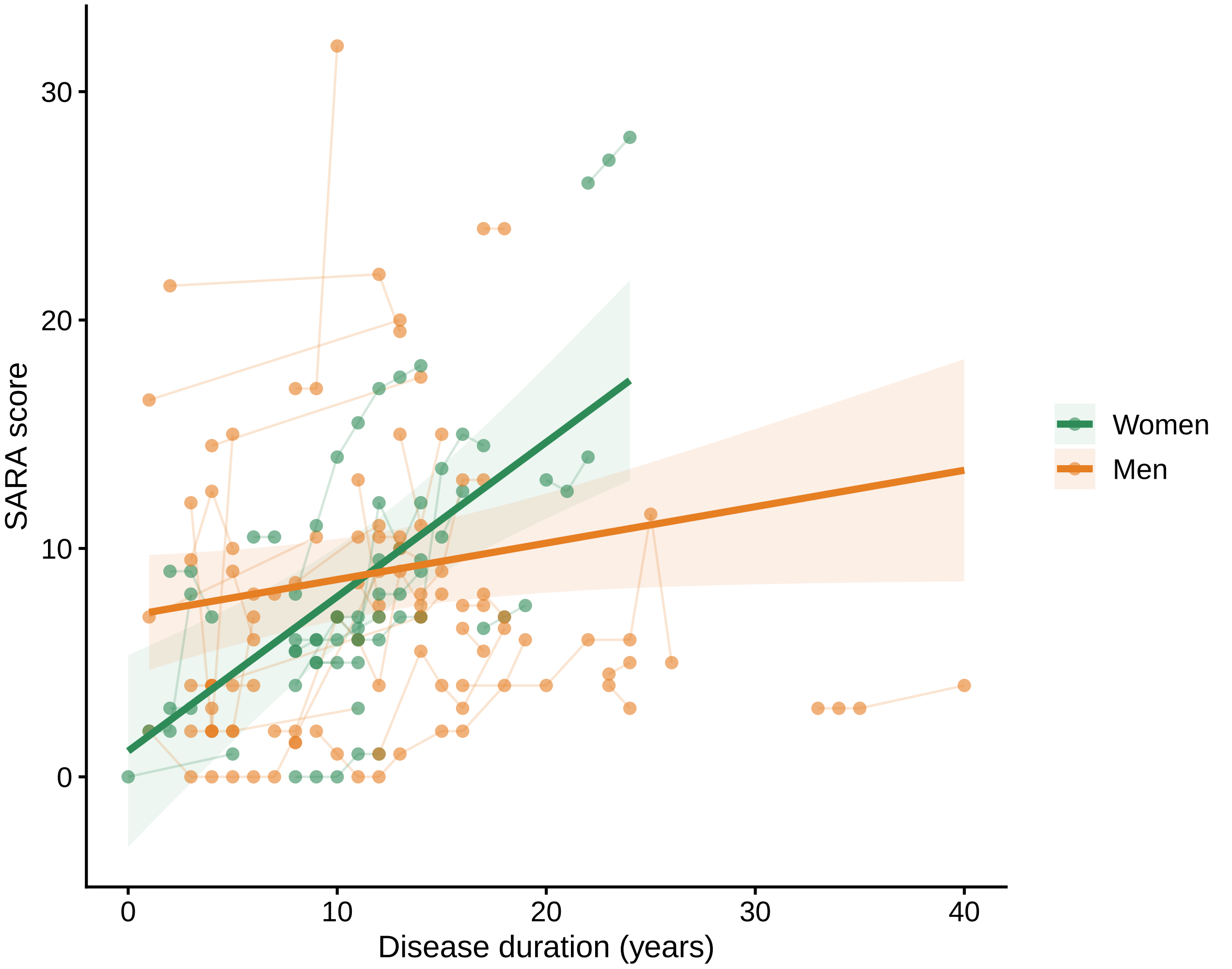
