## Supplementary Table 1 for "A Large Spanish Cohort Study Defines SCA27B Distinct Clinical Phenotype and its Longitudinal Progression"

| **Supplementary Table 1. Previous genetic studies** | | | |
| --- | --- | --- | --- |
| **Group** | **Molecular analysis** | **Negative** | **Not performed/ Not available** |
| **ILOCA**  **n=283** | Friedreich ataxia | 133 | 150 |
|  | FXTAS | 107 | 176 |
|  | SCA1 | 205 | 78 |
|  | SCA2 | 206 | 77 |
|  | SCA3 | 205 | 78 |
|  | SCA6 | 204 | 79 |
|  | SCA7 | 205 | 78 |
|  | SCA12 | 147 | 136 |
|  | SCA17 | 186 | 97 |
|  | SCA36 | 63 | 220 |
|  | DRPLA | 183 | 100 |
|  | RFC1 | 83 | 200 |
|  | NGS ataxia panel | 96 | 187 |
|  | WES | 31 | 252 |
|  | WGS | 3 | 280 |
| **SCA27B**  **n=217** | Friedreich ataxia | 77 | 140 |
|  | FXTAS | 66 | 151 |
|  | SCA1 | 127 | 90 |
|  | SCA2 | 127 | 90 |
|  | SCA3 | 120 | 97 |
|  | SCA6 | 128 | 89 |
|  | SCA7 | 124 | 93 |
|  | SCA12 | 85 | 132 |
|  | SCA17 | 113 | 104 |
|  | SCA36 | 57 | 160 |
|  | DRPLA | 112 | 105 |
|  | RFC1 | 72 | 145 |
|  | NGS ataxia panel | 64 | 153 |
|  | WES | 25 | 192 |
|  | WGS | 4 | 213 |
