## Supplementary Table 2 for "A Large Spanish Cohort Study Defines SCA27B Distinct Clinical Phenotype and its Longitudinal Progression"

| **Supplementary Table 2. Comparison of clinical characteristics between patients with monoallelic and biallelic expansions** | | | | |
| --- | --- | --- | --- | --- |
| **Parameter** | **Monoallelic n = 203** | **Biallelic   n = 14** | **P** | **P-FDR** |
| Age at onset**,** median [Q1-Q3]/N | 63 [52–69]/197 | 60 [55–64]/13 | 0.174 | 1.000 |
| Age at evaluation, median [Q1-Q3]/N | 74 [65–80]/195 | 74 [67.2–76.8]/12 | 0.630 | 1.000 |
| Disease duration at evaluation, median [Q1-Q3]/N | 10 [6–15]/189 | 11 [7.5–18.5]/11 | 0.496 | 1.000 |
| Women, n/N (%) | 87/203 (42.9%) | 9/14 (64.3%) | 0.164 | 1.000 |
| Episodic course, n/N (%) | 93/203 (45.8%) | 5/14 (35.7%) | 0.583 | 1.000 |
| Gait ataxia, n/N (%) | 168/180 (93.3%) | 12/12 (100%) | 1.000 | 1.000 |
| Impaired postural stability, n/N (%) | 65/166 (39.2%) | 4/11 (36.4%) | 1.000 | 1.000 |
| Impaired tandem gait, n/N (%) | 158/176 (89.8%) | 11/12 (91.7%) | 1.000 | 1.000 |
| Dysarthria, n/N (%) | 102/178 (57.3%) | 11/12 (91.7%) | 0.029 | 0.809 |
| Dysphagia, n/N (%) | 29/171 (17%) | 5/12 (41.7%) | 0.049 | 0.809 |
| Oculomotor disorders, n/N (%) | 161/194 (83%) | 9/12 (75%) | 0.444 | 1.000 |
| Downbeat nystagmus, n/N (%) | 86/129 (66.7%) | 5/6 (83.3%) | 0.663 | 1.000 |
| Finger-to-nose dysmetria | 116/180 (64.4%) | 7/12 (58.3%) | 0.759 | 1.000 |
| Heel-to-shin dysmetria | 123/178 (69.1%) | 6/12 (50%) | 0.205 | 1.000 |
| Parkinsonism | 15/176 (8.5%) | 1/11 (9.1%) | 1.000 | 1.000 |
| Tremor | 38/177 (21.5%) | 2/12 (16.7%) | 1.000 | 1.000 |
| Spasticity | 9/178 (5.1%) | 0/12 (0%) | 1.000 | 1.000 |
| Impaired light touch and pain sensation | 4/159 (2.5%) | 0/11 (0%) | 1.000 | 1.000 |
| Reduced vibration sense | 53/174 (30.5%) | 2/12 (16.7%) | 0.514 | 1.000 |
| Impaired joint position sense | 6/153 (3.9%) | 1/11 (9.1%) | 0.391 | 1.000 |
| Restless leg syndrome | 3/169 (1.8%) | 0/10 (0%) | 1.000 | 1.000 |
| Hyporeflexia | 32/177 (18.1%) | 2/12 (16.7%) | 1.000 | 1.000 |
| Hyperreflexia | 43/178 (24.2%) | 4/12 (33.3%) | 0.495 | 1.000 |
| Extensor plantar response | 7/173 (4%) | 1/12 (8.3%) | 0.422 | 1.000 |
| Weakness/atrophy | 6/177 (3.4%) | 1/12 (8.3%) | 0.373 | 1.000 |
| Vestibular areflexia |  |  | 0.883 | 1.000 |
| No vestibular areflexia | 89/120 (74.2%) | 3/4 (75%) |  |  |
| Positive HIT upon clinical examination | 21/120 (17.5%) | 0/4 (0%) |  |  |
| Video positive HIT | 10/120 (8.3%) | 1/4 (25%) |  |  |
| Cough | 10/174 (5.7%) | 0/12 (0%) | 1.0000 | 1.0000 |
| Dysautonomia | 25/170 (14.7%) | 1/12 (8.3%) | 1.0000 | 1.0000 |
| Cognitive decline |  |  | 0.9140 | 1.0000 |
| No cognitive decline | 146/172 (84.9%) | 11/13 (84.6%) |  |  |
| MCI | 23/172 (13.4%) | 1/13 (7.7%) |  |  |
| Dementia | 3/172 (1.7%) | 1/13 (7.7%) |  |  |
| Hearing loss | 19/174 (10.9%) | 1/11 (9.1%) | 1.0000 | 1.0000 |
| Headache | 11/162 (6.8%) | 0/11 (0%) | 1.0000 | 1.0000 |
| Epilepsy | 2/113 (1.8%) | 0/9 (0%) | 1.0000 | 1.0000 |
