## Supplementary Table 3 for "A Large Spanish Cohort Study Defines SCA27B Distinct Clinical Phenotype and its Longitudinal Progression"

| **Supplementary Table 3. Electroneurography and neuroimaging findings in patients with monoallelic and biallelic expansions** | | | | |
| --- | --- | --- | --- | --- |
| **Variable** | **Monoallelic n = 203** | **Biallelic   n = 14** | **P** | **P-FDR** |
| Cranial MRI, n/N (%) | 190/198 (96%) | 12/14 (85.7%) | 0.134 | 1 |
| Vermian atrophy, n/N (%) | 101/182 (55.5%) | 8/12 (66.7%) | 0.556 | 1 |
| Cerebellar hemispheric atrophy, n/N (%) | 74/175 (42.3%) | 8/12 (66.7%) | 0.134 | 1 |
| Leukoencephalopathy, n/N (%) | 21/167 (12.6%) | 1/10 (10%) | 1.000 | 1 |
| Cerebral atrophy, n/N (%) | 47/163 (28.8%) | 4/10 (40%) | 0.483 | 1 |
| Brainstem atrophy, n/N (%) | 5/161 (3.1%) | 0/10 (0%) | 1.000 | 1 |
| Brainstem signal abnormality, n/N (%) | 6/164 (3.7%) | 0/12 (0%) | 1.000 | 1 |
| Calcifications, n/N (%) | 1/160 (0.6%) | 0/10 (0%) | 1.000 | 1 |
| Iron deposition, n/N (%) | 2/158 (1.3%) | 0/10 (0%) | 1.000 | 1 |
| Spinal MRI, n/N (%) | 53/159 (33.3%) | 3/12 (25%) | 0.753 | 1 |
| Spinal cord atrophy, n/N (%) | 1/53 (1.9%) | 0/3 (0%) | 1.000 | 1 |
| Nerve conduction studies, n/N (%) | 94/164 (57.3%) | 7/12 (58.3%) | 1.000 | 1 |
| Normal sensory electroneurogram, n/N (%) | 85/94 (90.4%) | 6/7 (85.7%) | 0.530 | 1 |
| Sensory axonal neuropathy, n/N (%) | 8/94 (8.5%) | 1/7 (14.3%) | 0.491 | 1 |
| Normal motor electroneurogram, n/N (%) | 85/94 (90.4%) | 6/7 (85.7%) | 0.530 | 1 |
| Motor axonal neuropathy, n/N (%) | 7/94 (7.4%) | 0/7 (0%) | 1.000 | 1 |
